# From Local Inhibition to Distributed Motor Control in Aging

**DOI:** 10.64898/2026.08.03.26359573

**Authors:** Lea-Theresa Mais, Sebastian Dern, Nan Liu, Gereon R. Fink, Christian Grefkes, Caroline Tscherpel

**Author notes:** Corresponding author. Medical Faculty, Goethe University Frankfurt, and Department of Neurology, University Hospital Frankfurt, Frankfurt am Main, Germany, *E-Mail address:* (C. Tscherpel).

## Abstract

**Background:** Aging-related changes in motor performance are associated with a widespread reorganization of the motor network. Despite extensive research on altered brain activity patterns in aging, the underlying neurophysiological mechanisms driving these reorganizational changes remain incompletely understood, particularly regarding the role of inhibitory control.

**Methods:** We here combined transcranial magnetic stimulation (TMS) and electroencephalography (EEG) to examine aging-related alterations in local excitability, oscillatory dynamics, and neural coupling in the motor system. Brain responsivity was probed by applying single-pulse TMS to the primary motor cortex (M1) in younger and older subjects at rest.

**Results:** Our findings indicated reduced cortical excitability in the stimulated M1 of older participants, consistent with an age-related reduction of GABAergic inhibitory control. Phase-locking analyses revealed reduced intra- and interhemispheric coupling in lower frequency bands, suggesting decreased inhibitory output from the stimulated M1. Moreover, older individuals demonstrated less localized event-related desynchronization (ERD) with greater power decreases in the prefrontal cortex contralateral to the stimulation site, indicating enhanced prefrontal involvement in the aging motor system.

**Conclusions:** Together, these findings underline the relevance of alterations in inhibitory processes in the motor network in aging and point to a shift from an automatic, locally driven towards a broader, putatively more cognitively controlled sensorimotor processing in older adults.

## Introduction

Aging is characteristically accompanied by deterioration of motor performance [1]. However, aging also involves widespread changes in cortical and subcortical circuits [2,3]. Findings from task-based functional magnetic resonance imaging (fMRI) have shaped frameworks of age-related reorganization, such as the posterior-anterior shift in aging (PASA) and the hemispheric asymmetry reduction in older adults (HAROLD) model [4,5]. While such observations are frequently interpreted as compensatory to counterweigh age-related degeneration, the dedifferentiation hypothesis instead proposes that changes in activity patterns indicate a loss of neural specialization associated with altered neurotransmission [6]. However, recent literature rather unifies both ideas in the compensation-related utilization of neural circuits (CRUNCH) hypothesis, proposing that older individuals engage additional neural resources at lower task demands, which eventually reach a ceiling with increasing task complexity, thereby reflecting both compensatory mechanisms and neural limitations [2,7,8].

Despite extensive work on altered brain activity patterns in aging, the underlying neural mechanisms driving these reorganizational changes remain incompletely understood. Besides age-related changes in activity patterns, fMRI studies have reported increased connectivity during rest and motor tasks in older participants [3,9,10]. These findings of altered network integration are supported by EEG research [11,12,13], which, due to its high temporal resolution, is particularly well-suited to capture the fast and dynamic processes underlying motor control. Notably, besides a general increase in connectivity, Rosjat and colleagues [14] reported increased motor coupling with frontal regions, possibly reflecting enhanced top-down modulation.

EEG also enables the characterization of neural oscillations, which have been shown to be profoundly relevant in sensorimotor and cognitive functions [15]. During motor tasks, aging has been associated with a greater and more bilateral power decrease in the alpha and beta frequency bands [16]. Moreover, pharmacological studies have linked these changes to GABAergic inhibition, with experimentally increased cortical GABA levels resulting in larger desynchronization amplitudes, indicating that older adults recruit broader inhibitory neural resources [16,17]. At rest, aging has been linked to a slowing of oscillatory activity, characterized by reduced alpha power and an increase in the theta and delta frequency bands [18]. In addition, coupling in the motor system was found to be predominantly altered in the slow frequency bands, again underlining the crucial role of slow oscillations in aging [14,19,20]. Notably, delta and theta oscillations have been associated with GABA-mediated inhibitory processes [21,22,23], supporting the link between age-related changes and altered GABAergic inhibitory control.

Accordingly, previous studies suggested that GABA may be a factor contributing to age-related functional deterioration [24]: Animal studies found age-related changes in the GABA signaling system across species, including decreased GABA_A_ receptor subunit expression and reduced GABAergic neuron density [25,26,27,28]. Furthermore, human studies using magnetic resonance spectroscopy (MRS) reported an age-related decrease across various brain areas, including primary and secondary motor areas [29,30,31,32,33], with some linking lower GABA levels to reduced behavioral performance. However, mixed results have been observed in regions of interest, such as the frontal and sensorimotor cortex, with both reduced [29,34,35] and unchanged [34,36,37] levels of GABA in aging. Consequently, whether and how changes in GABAergic inhibitory processes contribute to alterations of motor control during aging remains under debate.

In this context, transcranial magnetic stimulation (TMS) combined with EEG (TMS-EEG) represents a non-invasive perturb-and-measure approach that offers a direct assessment of cortical excitability and inhibition. It combines temporally and spatially precise perturbations to systematically assess changes in local excitability, oscillatory dynamics, and signal propagation [38,39]. Moreover, TMS-evoked potentials (TEPs) have been linked to distinct excitatory and inhibitory mechanisms, such as GABAergic inhibition [40,41]. TMS-EEG studies examining motor cortical excitability in aging remain scarce and have primarily focused on age-related changes in TMS-evoked potentials (TEPs), resulting in heterogeneous findings [42,43,44]. However, a comprehensive characterization of changes in oscillatory dynamics and coupling, along with differences in excitability of evoked potentials, might provide a more detailed understanding of how age-related changes in inhibitory processes contribute to observed changes in activity patterns in aging.

To address these questions, we applied neuronavigated TMS to the primary motor cortex (M1) combined with high-density EEG to probe local cortical excitability and network dynamics in both young and older individuals. Analyses comprised three complementary domains: (i) local excitability, (ii) oscillatory dynamics, and (iii) neural coupling. These domains can provide distinct yet interrelated insights into alterations of inhibitory mechanisms at the regional and network level following non-invasive cortical stimulation. Based on prior evidence of an age-related reduction in GABAergic activity [29] and findings linking TMS-evoked potentials to the excitation-inhibition balance mediated by GABA and glutamate circuits [41], we hypothesized a reduction in local TMS-evoked activity within the stimulated motor area in older participants, reflecting a decrease in inhibitory activity. In line with previous findings of increased frontal cortex engagement (i.e., PASA), we further anticipated a similar upregulation of frontal involvement, particularly in the lower frequency bands indexing age-associated changes in inhibitory processes.

## Materials and Methods

### Participants

Our study sample consisted of 32 healthy individuals: 18 younger participants (3 females; mean age of 24.4±3.2 standard deviation (SD) years, range: 20–30 years; 17 right-handed according to the Edinburgh Handedness Inventory (EHI)) and 14 older participants (2 females; 65.5±6.2 years, range: 51-77 years; 13 right-handed) were recruited from our database. The following criteria led to exclusion of subjects: (i) any contraindication to TMS (e.g., epilepsy, pacemaker) [45], (ii) history of neurological, psychiatric, orthopedic, or rheumatic diseases. All individuals gave informed written consent before participating in the study. The study was approved by the ethics committee of the medical faculty at the University of Cologne (file no. 17-244) and carried out in accordance with the Declaration of Helsinki. Notably, parts of the original data, i.e., data from the 14 older participants, were previously published in Tscherpel et al. [46,47]. However, as the former work focused on reorganization and recovery after stroke, and older subjects were included only as an age-matched control group to emphasize the specificity of findings after stroke, there is no overlap between the analyses presented here and those published previously. Hence, all analyses reported are novel.

### Experimental procedure and TMS

Before starting with the TMS-EEG recordings, maximum grip force was assessed in three consecutive trials separately for each hand using a vigorimeter (KLS Martin Group). Participants were stimulated with a Magstim Super Rapid^2^ stimulator (The Magstim Co. Ltd.) equipped with a Magstim 70mm figure-of-eight Alpha Film Coil. This setup was complemented by a Brainsight neuronavigation system (V.2.0.7; Rogue Research Ltd.) to ensure precise coil positioning throughout the experiment. The ‘motor hotspot’ and resting motor threshold (RMT) were defined according to standard procedures described in detail in previous publications [46,47] (see Supplementary Material I for further information).

### TMS-EEG recordings

We used a TMS-compatible 64-channel EEG system (BrainAmp DC, BrainProducts). In accordance with the standard layout and the international 10-20 system, 62 TMS-compatible Ag/AgCl sintered ring electrodes were positioned on an elastic electrode cap (EasyCap-Fast’n Easy 64Ch). For the experiment, we stimulated the participant’s individual motor hotspot with a suprathreshold stimulation intensity of 110% RMT to ensure a sufficient signal-to-noise ratio [48,49,50,51]. 100 trials of single TMS pulses with a randomly jittered inter-trial interval of 6.5-8.0s were applied. Procedures and hardware settings previously described were used to ensure high-quality recordings and minimize artifacts (see [47] and Supplementary Material II).

### Data analysis

Matlab R2023a (The MathWorks, Massachusetts, USA) and in-house scripts based on functions of the open-source toolbox EEGLAB (https://sccn.ucsd.edu/eeglab/) [52] were used for data preprocessing and analyses. Data were preprocessed according to procedures described in detail in previous publications [46,47] (see Supplementary Material III).

Cluster-based permutation analysis and LMFP of identified regions of interest

First, we investigated TMS-evoked EEG characteristics at each electrode using a non-parametric cluster-based permutation analysis, which allows comparisons between age groups and time points with robust protection against multiple comparison errors [53] (for further details see Supplementary Material).

Based on the results of the cluster-based permutation test, we identified two areas for further analyses: a centroparietal region of interest (ROI), comprising electrodes C3 and C5 above the ipsilateral motor cortex, as well as a frontal ROI contralateral to the stimulation site, including electrodes F2 and F4.

To explore age-related changes within these ROIs, we calculated the local mean field power (LMFP). The LMFP provides a global index of EEG activity over time and is calculated as the root-mean-squared value of the data across the channels of interest [54,55]. Based on the result of the cluster-based permutation analysis, we examined the area-under-the-curve (AUC) within three distinct time windows representing the early phase after the stimulus onset (20-100ms), a time window between 100ms and 200ms, and a later post-stimulus stage (200-350ms).

### Time-frequency analysis

To characterize age-related alterations in neural oscillations, we transformed the TMS-EEG data into the time-frequency domain using the EEGLAB function ‘newtimef’ [52] with a Morlet wavelet transform (1-7 cycles, increasing with frequency). We then computed the event-related spectral perturbation (ERSP) within the 1-30Hz frequency range to examine spectral features. We performed absolute spectra normalization at the individual-trial level [56], then averaged the ERSP across trials and applied a pre-stimulus baseline correction (−500ms to −100ms). Bootstrap-based statistics (number of permutations=500, alpha-level<0.05) were used to identify significant ERSP values relative to baseline. To investigate differences in event-related synchronization (ERS) and event-related desynchronization (ERD) after motor cortex stimulation, significant ERSP values for the frontal and centroparietal ROIs within the 8-30Hz frequency range were averaged within two distinct time windows: 0-200ms and 200-350ms post-stimulus [49].

#### Phase-locking value

Finally, to investigate age-related differences in TMS-evoked neuronal coupling, we calculated the phase locking value (PLV) between the stimulation site (C3) and all other electrodes using the open-source MATLAB-based toolbox Brainstorm (http://neuroimage.usc.edu/brainstorm). PLV makes use of the phase difference between two time series and is robust against fluctuations in signal amplitude [57,58]. Before PLV analyses, we computed current source density (CSD) estimates using spherical splines [59] with the CSD toolbox [60] to reduce volume-conduction effects and, thereby, improve spatial resolution. We excluded edge electrodes, leaving 39 electrodes for further analyses. We next examined the PLV from C3 to four regions of interest: an ipsilateral centroparietal region surrounding C3 (CP3, C5, CP5), a contralateral centroparietal region (C4, CP4, C6, CP6), representing ipsi- and contralateral motor areas, an ipsilateral frontal region (F1, F3), and a contralateral frontal region (F2, F4). Furthermore, we averaged the PLV values over four frequency bands: δ (1-4Hz), θ (4-8Hz), α (8-12Hz), and β (13-30Hz).

### Statistical analysis

We used SPSS (Statistical Package for the Social Science, version 29, IBM) for statistical analyses. To decide between parametric or non-parametric statistics, we first confirmed normal distribution of our data using Kolmogorov-Smirnov tests. Accordingly, group differences between younger and older participants were evaluated using either independent two-sided t-tests or non-parametric Mann-Whitney U-tests (p<0.05). To additionally compare between regions of interest and frequency bands, we implemented repeated-measures analyses of variance (ANOVAs) testing for the interaction of the factor GROUP (two groups: younger; older) x factor LOCALIZATION (two regions: centroparietal; frontal) for ERSP analyses and factor GROUP (two groups: younger; older) x factor LOCALIZATION (four regions: ipsilateral centroparietal; contralateral centroparietal; ipsilateral frontal; contralateral frontal) x factor FREQUENCY (four frequency bands: δ; θ; α; β) for PLV analyses. To further investigate the contribution of neurophysiological and behavioral measures to age-related variance, linear regression analyses were conducted with age as the dependent variable. LMFP of the centroparietal ROI, ERD of centroparietal and frontal ROIs, PLV values in centroparietal ROIs (δ, θ), and grip force were included as independent variables. For correlation analyses, we used Pearson’s r in normally distributed data and Spearman’s rho for non-parametric statistics. Multiple comparisons in post-hoc tests and correlation analyses were corrected for false discovery rate (FDR-correction).

### Data availability

The data underlying the findings of this study, along with Matlab scripts used for analysis, can be obtained from the corresponding author upon reasonable request.

## Results

### TMS Parameters

There were no significant differences in the resting motor threshold (rMT) between younger (54.44 ± 7.16% maximum stimulator output (MSO)) and older individuals (55.07 ± 11.61% MSO) (p=0.87, Z=0.19, Mann-Whitney U-Test). Hence, age-related differences in TMS-EEG parameters were not driven by differences in stimulation intensities.

### Age-related alterations in TMS-evoked potentials after motor cortex stimulation

The cluster-based permutation analyses revealed two significant clusters of differential TMS-EEG responses (Fig. 1). One cluster, ranging from 90ms to 200ms post-stimulus and representing more negative potentials in younger participants, involved electrodes in the ipsilateral centroparietal area surrounding the stimulation site (p=0.008). Additionally, we found a second significant cluster from 60ms to 180ms with more negative potentials in older participants, comprising electrodes in frontal areas contralateral to the stimulation site (p=0.012). See Supplementary Material IV for further details on significant clusters.

**Figure 1.**
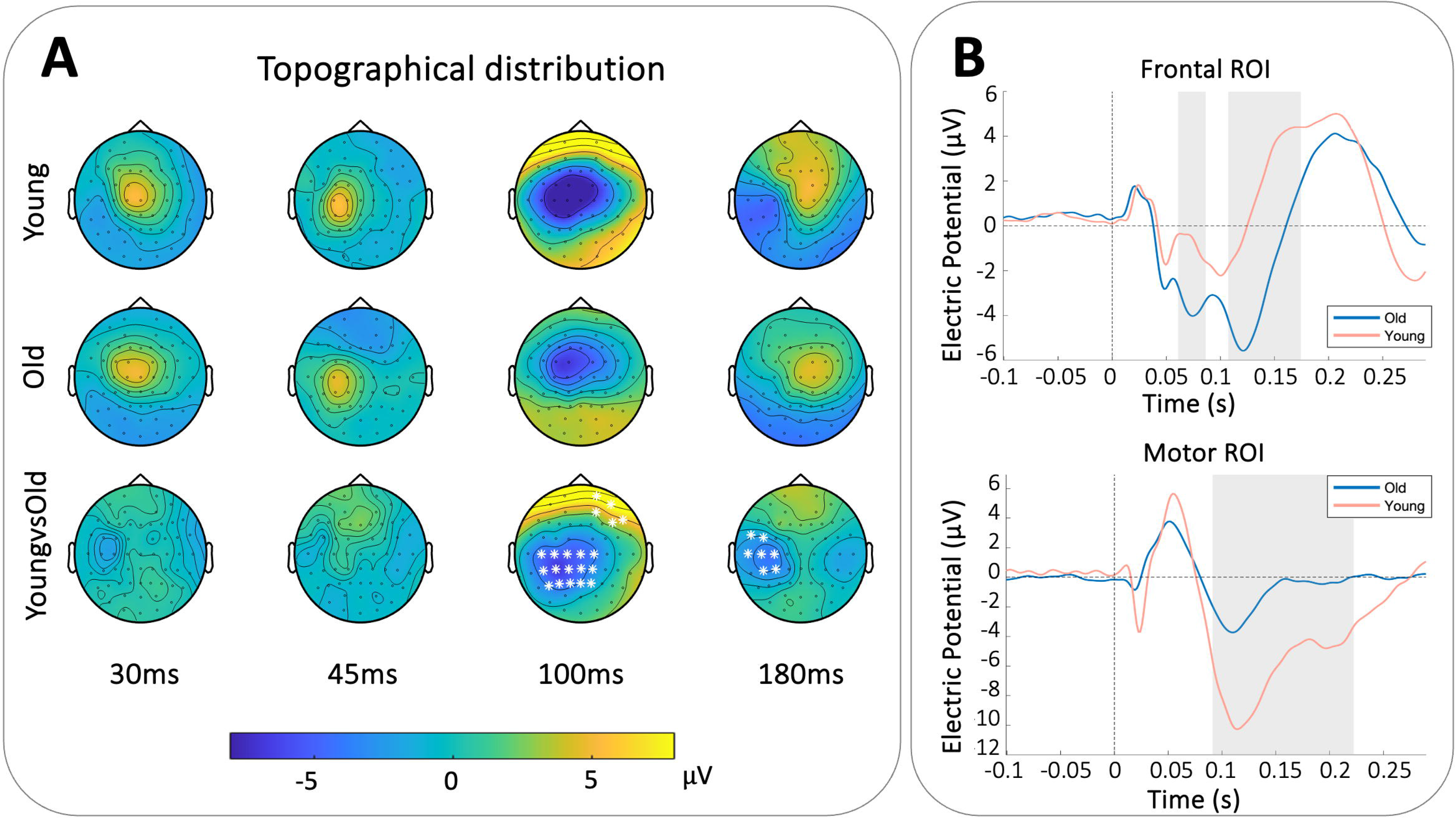
Comparison of TMS-evoked potentials between younger and older adults. (A) Topographical distribution of TMS-evoked activity in μV at 30, 45, 100, and 180 ms after TMS applied to the primary motor cortex. The bottom row depicts difference values between younger and older adults, with white stars indicating electrodes contributing to significant clusters in the cluster-based permutation test. (B) ERP-plots with mean electric potential in C3C5 and F2F4 regions of interest. The gray area masks the time frame of a significant difference between age groups. The vertical dashed line indicates the timing of the TMS pulse.

Comparison of the LMFP in the ipsilateral centroparietal ROI (C3C5) revealed a significantly reduced TMS-evoked response in older adults in all time windows derived by the results of the cluster-based permutation analysis (20-100ms: p=0.018, Z=2.36; 100-200ms: p<0.001, Z=3.84; 200-350ms: p=0.002, Z=3.27, Mann-Whitney U-tests, FDR-corrected), indicating decreased cortical responsivity in older compared to younger individuals. In contrast, no significant difference between groups was observed in the contralateral frontal ROI (F2F4) in any of the time windows (20-100ms: p=0.512, Z=-0.68; 100-200ms: p=0.151, t_(30.0)_=-1.69; 200-350ms: p=0.151, Z=1.67, FDR-corrected).

### Differences in event-related synchronization and desynchronization

Regarding TMS-evoked EEG-responses in the time-frequency domain (ERSP), both younger and older participants showed a broadband power increase within 200ms post-stimulation (Fig. 2). Accordingly, a repeated-measures ANOVA for the 0-200ms time window revealed a significant main effect of the factor LOCALIZATION with higher power in the ipsilateral centroparietal compared to the contralateral frontal region (p<0.001, F_(1.0,30.0)_=14.42), but neither a significant main effect for GROUP (p=0.116, F_(1.0,30.0)_=2.62) nor an interaction between GROUP x LOCALIZATION (p=0.094, F_(1.0,30.0)_=3.0). However, following the initial increase in power, a decrease in power, i.e., ERD, in the 8-30Hz frequency range was observed after 200ms post-stimulus. Here, a repeated measures ANOVA for the 200-350ms time window revealed a significant interaction effect for GROUP x LOCALIZATION (p=0.014, F_(1.0,30.0)_=6.75), indicating age- and region-dependent differences in desynchronization. We also found a significant main effect of LOCALIZATION (p=0.006, F_(1.0,30.0)_=8.89) but no main effect of GROUP (p=0.239, F_(1.0,30.0)_=1.44). Post-hoc tests revealed significantly greater desynchronization in older than in younger participants in the contralateral frontal regions (p=0.039, t_(30.0)_=-2.47) but not in the ipsilateral centroparietal region (p=0.957, t_(30.0)_=-0.08). Interestingly, while in younger participants desynchronization was significantly reduced in the frontal region compared to the centroparietal area (p=0.01, t_(17.0)_=-3.71), this centrofrontal gradient was not evident in older participants with nearly similar desynchronization values in both regions of interest (p=0.957, t_(13.0)_=-0.32, two-sided t-test, FDR-corrected), indicating a more widespread desynchronization across the cortex in older individuals compared to a rather regionally confined pattern in younger participants.

**Figure 2.**
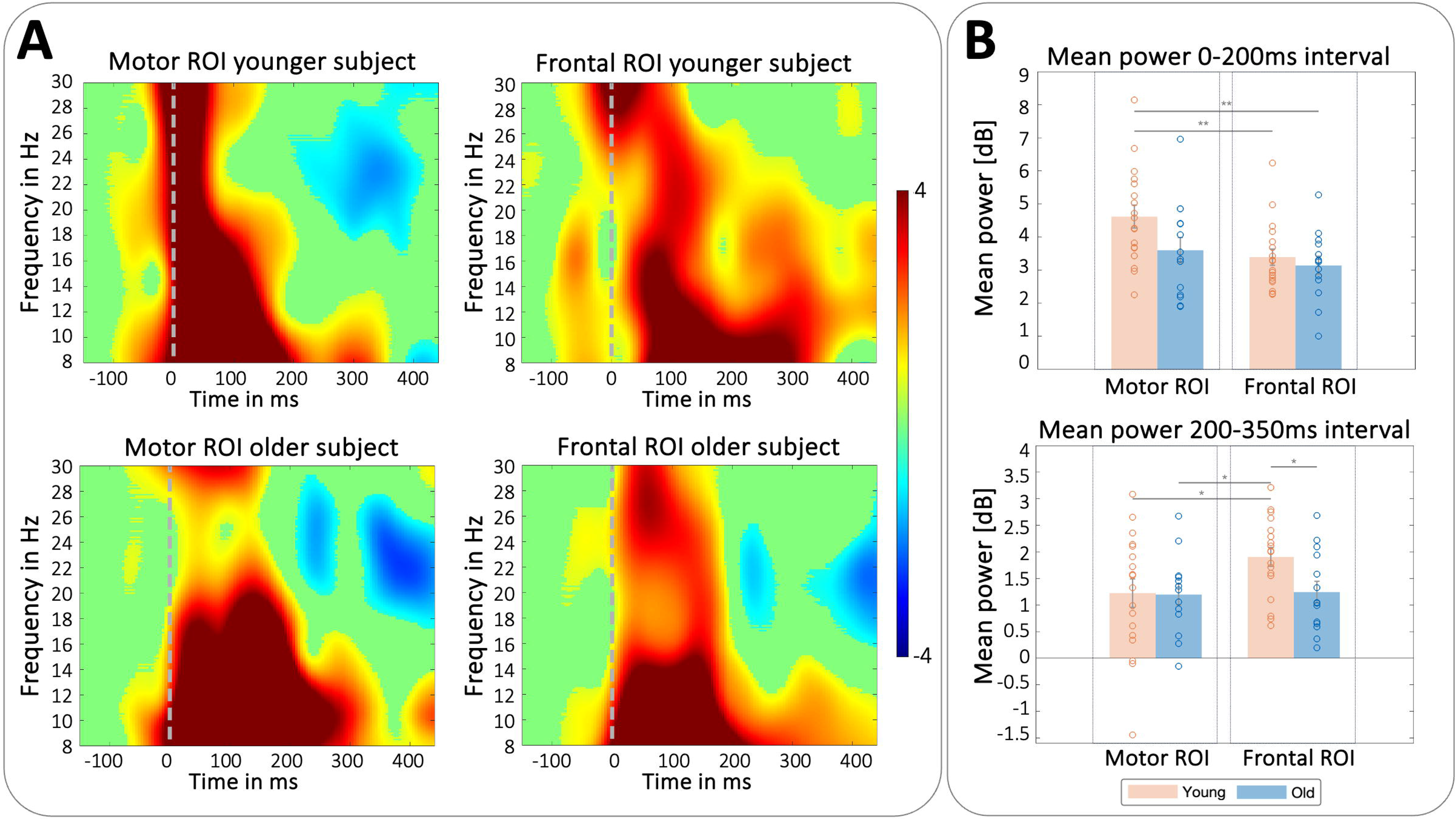
Time-frequency analyses of alpha and beta frequency bands. (A) ERSP plots in one younger and one older subject showing the development of frequency band power over time. Colourbar depicts power in dB. The vertical dashed line indicates the timing of the TMS pulse. (B) For each time interval analyzed, the bar chart shows mean power values in the regions of interest compared between younger and older adults. Error bars indicate the standard error. (* p < 0.05; ** p < 0.01)

### Age-related reduction in TMS-evoked neuronal coupling

To explore age-related alterations in TMS-evoked connectivity, we examined the phase-locking values (PLVs) between the stimulation site (C3) and four regions of interest: ipsi- and contralateral centroparietal regions (CP3, C5, CP5 and C4, CP4, C6, CP6) and ipsi- and contralateral frontal areas (F1, F3 and F2, F4), selected to capture both intra- and interhemispheric connectivity. The analyses revealed a significant interaction effect for GROUP x LOCALIZATION x FREQUENCY (p=0.022, F_(5.31,159.2)_=2.67), indicating that differences in PLV depend on age, region, and frequency band (Fig. 3). Accordingly, post-hoc tests considering coupling within the ipsilateral centroparietal region under the stimulation site revealed that PLV differed significantly in the lower δ- and θ-frequency bands between age groups with reduced phase-locking values in older subjects (δ: p=0.009, t_(30.0)_=-3.34; θ: p=0.01, t_(30.0)_=-2.99; α: p=0.896, t_(30.0)_=-0.13; β: p=0.817, t_(30.0)_=-0.51). This indicates a reduced integration of the ipsilateral motor area in older participants, particularly in lower frequency bands. Moreover, we further found significant differences in the δ- and θ-frequency for the coupling between the site of stimulation and the contralateral centroparietal region between younger and older adults (δ: p=0.033, t_(30.0)_=-2.48; θ: p=0.033, t_(30.0)_=-2.36; α: p=0.245, t_(30.0)_=-1.19; β: p=0.033, t_(30.0)_=-2.53, two-sided t-tests, FDR-corrected). Again, we observed reduced interhemispheric coupling between bilateral motor areas in older participants, further underlining a decline of TMS-evoked neuronal coupling with age. Post-hoc comparisons of coupling from the stimulation site to bilateral frontal ROIs revealed no significant differences between younger and older individuals.

**Figure 3.**
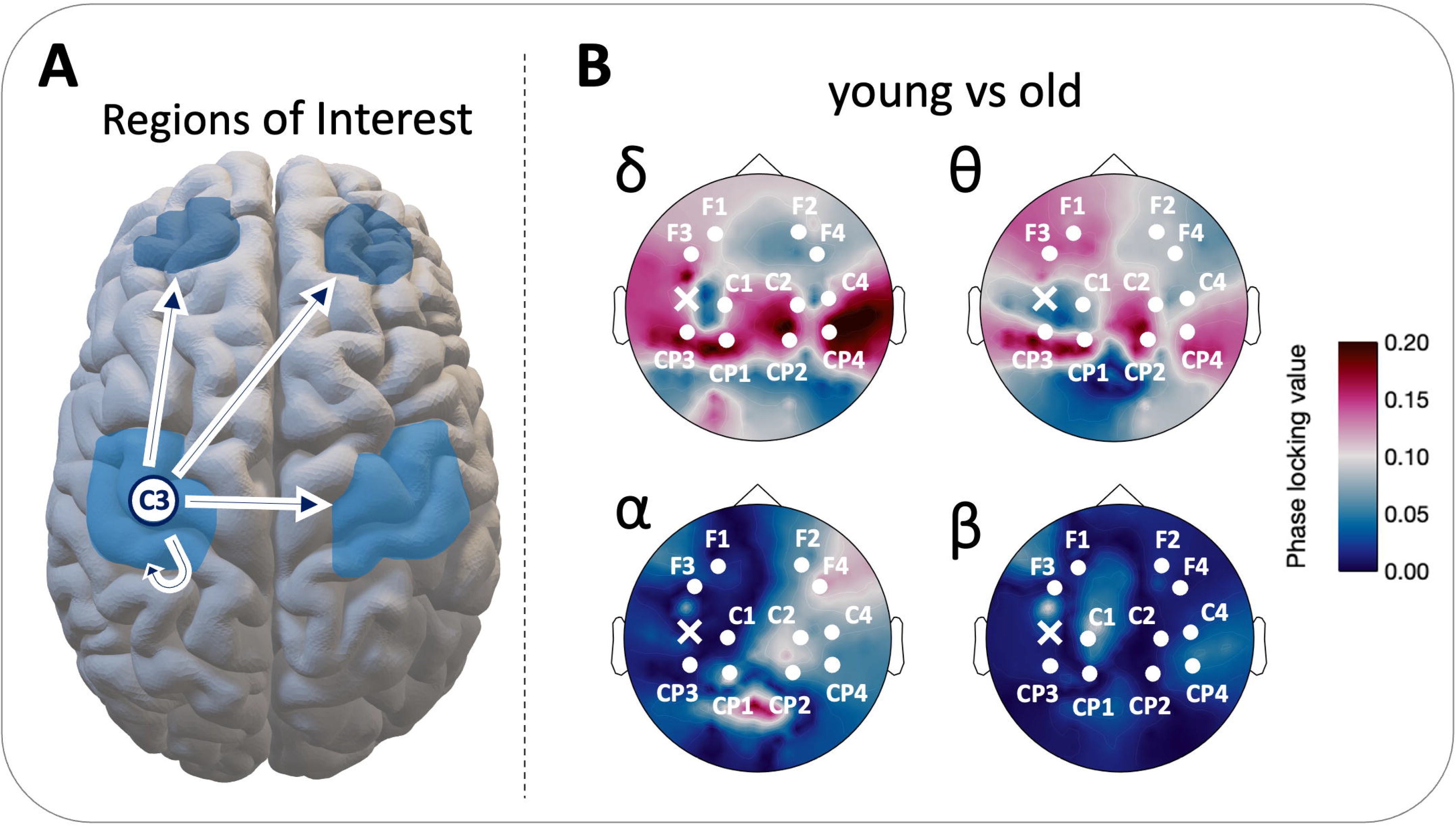
Connectivity of channel C3 with remaining channels. (A) Depicted are the ROIs for the phase-locking value analyses, with arrows demonstrating which coupling was assessed. (B) Difference in phase-locking values between younger and older subjects, with higher values reflecting greater phase locking in younger subjects. Phase-locking values are averaged over frequency bands (δ: 1-4 Hz; θ: 4-8 Hz; α: 8-12 Hz; β: 13-30 Hz).

### Relationship between age, TMS-EEG markers, and motor output

We further examined the relationship between age, TMS-EEG measures, and motor output using a stepwise multiple linear regression. The analysis aimed to investigate the contribution of (i) LMFP, (ii) ERD, (iii) PLV, and (iv) grip force to the variance in age. Among these, the LMFP of the centroparietal ROI during the 100-200ms post-stimulus window, along with ERD both in the centroparietal and frontal ROIs, were the strongest predictors of age (R^2^=0.57, adjusted R^2^=0.52, F_(3,28)_=12.33, p<0.001). The LMFP (100-200ms) accounted for 29% of unique variance, while the combination of LMFP and ERD in the centroparietal region explained 43% of variance. These results suggest that aging impacts neurophysiological measures such as LMFP and ERD, particularly in the stimulated motor cortex. Considering the entire study sample, correlation analysis between the LMFP of the centroparietal region during the 100-200ms post-stimulus window revealed a significant decrease of the LMFP with increasing age (p<0.001, rho=-0.71, Spearman correlation coefficient, FDR-corrected), suggesting that higher age is associated with a decrease in excitability of the ipsilateral centroparietal region (Fig. 4). However, when analyzed separately, correlations within both older and younger groups were not significant. Moreover, none of the other correlational analyses yielded significant results.

**Figure 4.**
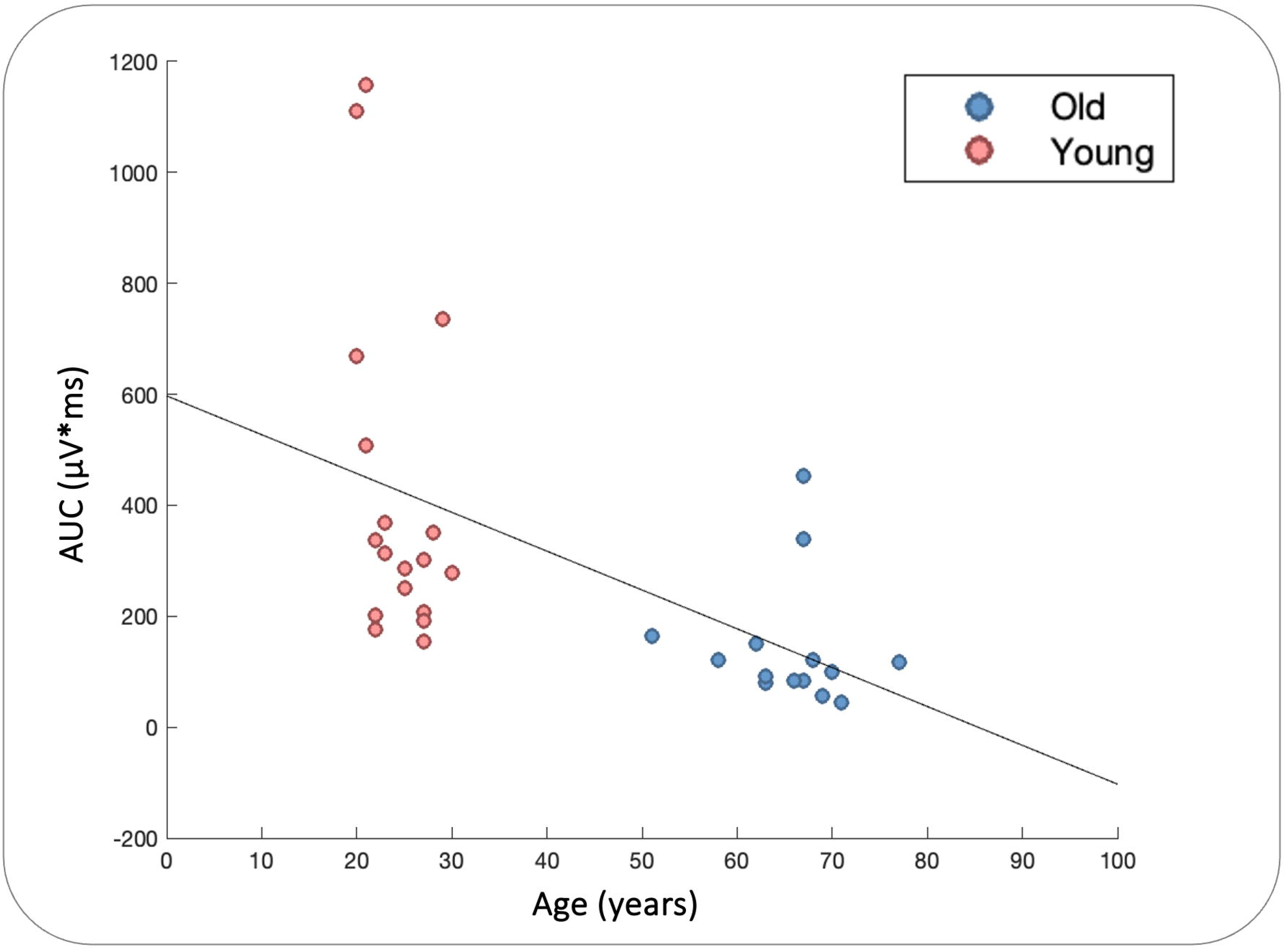
Relationship between age and LMFP in the motor ROI. Depicted is the correlation between age and the area under the curve (AUC) of the LMFP in the 100-200ms time window in the motor ROI.

## Discussion

We here used TMS-EEG to investigate age-related changes in cortical dynamics of the motor system. By characterizing cortical responsivity, oscillatory dynamics, and neural coupling during TMS-induced activation of the primary motor cortex, we demonstrate that aging is consistently accompanied by alterations across temporal, spectral, and network domains. Specifically, older participants showed reduced cortical reactivity of the primary motor cortex, attenuated low-frequency coupling between bilateral motor regions, and enhanced recruitment of the prefrontal cortex. These findings align with models of age-related functional adaptation, with hampered interhemispheric communication potentially underlying the reduced hemispherical lateralization described in the HAROLD model, a frontal shift in oscillatory dynamics reflecting the PASA pattern, and enhanced prefrontal cortex engagement suggesting compensatory mechanisms in line with the CRUNCH framework. As our data may link these age-related changes to GABAergic inhibitory processes, our findings suggest that altered inhibition may represent a common factor underlying reduced specialization and compensatory recruitment in the aging motor system [24,61].

### Age-related decline of the inhibitory response in M1

The TMS-evoked EEG response reflects a sequence of cortical events ranging from the immediate reactivity of local neuron populations to later large-scale network interactions [38,39]. Although the precise neural mechanisms underlying the TMS-evoked cortical response are not fully understood yet, converging evidence indicates that their magnitude and temporal dynamics are shaped by the balance between excitatory and inhibitory processes [41,61,62]. In particular, pharmacological TMS-EEG studies have shown that the modulation of GABAergic receptor activity alters the amplitude of TMS-evoked potentials, suggesting that GABAergic inhibition plays a central role in shaping the cortical response following stimulation [61,63,64].

In the present study, older participants exhibited a reduced TMS-EEG response within the stimulated motor cortex across all examined time windows, which based on the aforementioned knowledge may reflect an age-related attenuation of inhibitory regulation. Consistent with this interpretation, several lines of evidence indicate that the GABAergic system undergoes structural and functional alterations during aging [24,65,66]. At the cellular level, studies across species have reported reductions in GABAergic neuron density as well as alterations in inhibitory synaptic signaling within cortical circuits [27,28,65,67,68]. Changes have also been observed at the receptor level with decreased GABA receptor binding density and age-related alterations in receptor subunit composition, potentially affecting the efficacy of inhibitory neurotransmission [25,65,69,70,71,72,73]. In addition, in-vivo evidence for altered GABAergic inhibition comes from MRS studies reporting reduced cortical GABA concentrations in older adults across several brain regions, including the sensorimotor cortex [29,34]. Together, these findings suggest that aging is accompanied by alterations of GABAergic inhibitory neurotransmission, which may contribute to the difference in signal processing observed here. To put it in other words, the finding of a decreased TMS-EEG response in the primary motor cortex might therefore reflect a weakening of GABAergic inhibitory processes in the aging brain. In turn, such changes in inhibitory control may affect the efficiency of motor processing and could also provide a neurophysiological basis for altered network interactions.

### Reduced low-frequency coupling in the aging motor cortices

Alterations in network dynamics might also underlie the reduced functional hemispheric asymmetry observed in the aging brain [74]. The HAROLD model, initially framed for aging processes during cognitive tasks, describes the change from a laterally confined cortical activity observed in younger adults towards a more widespread brain activation pattern in older participants [4]. It has been interpreted as either a compensatory mechanism or dedifferentiation, reflecting a reduced ability to selectively recruit task-relevant regions. Prior research has suggested that unspecific disinhibition, i.e., reduced interhemispheric inhibition, might contribute to increased bilateral activity and, thereby, to the loss of lateralization in motor control [74,75,76,77,78].

In line with this notion, our results revealed reduced coupling between the stimulation site and both ipsilateral and contralateral motor areas, particularly within the delta and theta frequency ranges. Of note, phase-locking in lower frequency bands has been linked to critical aspects of motor control, including stimulus processing, movement initiation, and execution [14,19,79]. Previous studies have shown that interregional coupling changes significantly with aging [12,80]. Older individuals demonstrated not only reduced phase-locking within the core motor network but also additional intra- and interhemispheric connections, particularly involving premotor and frontal regions [12,14]. In line with that, we here found reduced low-frequency coupling within and between both motor areas in aging, indicating that, in addition to the alterations of local responsivity of the stimulated motor cortex, aging is also associated with changes of intra- and interhemispheric network dynamics.

Furthermore, delta and theta oscillations have also been linked to inhibitory control [81,82,83,84], suggesting that the observed changes are not only a consequence of age-related changes in local inhibition but also due to alterations of inhibitory interactions between cortical regions during aging. This interpretation is supported by previous TMS-EMG studies that have attributed reduced interhemispheric connectivity to a decline in transcallosal inhibitory drive between motor cortices [74,76,78]. The reduced phase-locking within and between motor cortices observed here may therefore indicate diminished inhibitory signaling in aging. In particular, slow oscillations are thought to be generated by thalamocortical circuits involving GABAergic mechanisms [21,22,23]. Therefore, age-related alterations of GABAergic modulation could have contributed to the reduced low-frequency coupling observed in our older participants. Consequently, our results support the notion that transcallosal inhibition is altered during aging and may contribute to the less lateralized activity patterns described by HAROLD.

### Increased prefrontal cortex involvement in oscillatory dynamics

To counteract age-related processing alterations, such as reductions in local inhibitory control and interhemispheric coupling, the aging brain may recruit additional neural resources [8,85], as proposed in frameworks such as PASA and CRUNCH. PASA describes an anterior shift with increased activity in more frontal regions, whereas CRUNCH posits that such additional recruitment might reflect a compensatory mechanism up to a ceiling point, beyond which older adults reach their neural limits [5,8]. Similar to findings from fMRI, studies investigating motor-related oscillatory dynamics by means of EEG have likewise reported a more widespread recruitment of cortical regions in older adults [16]. In particular, movement-related desynchronization in the alpha and beta frequency bands appears less localized in aging, including more pronounced decreases in frontal areas [16,85]. While alpha and beta ERD during motor execution is more generally interpreted as a reflection of the cortical regions’ activation involved in generating motor commands and processing cognitive and sensory information [16], desynchronization following TMS has been more specifically attributed to proprioceptive feedback processing arising from activation of the target muscle [49].

In our TMS-EEG data, older participants showed a lack of focal motor cortex desynchronization in the alpha and beta frequency bands commonly observed in younger participants [49], with a more widespread pattern including enhanced desynchronization in the contralateral prefrontal cortex. Previous task-based studies have linked increased frontal engagement to better motor performance in older adults, supporting the notion that enhanced recruitment serves to counteract functional decline [3,86,87]. Given the role of the prefrontal cortex in movement monitoring and cognitive control of actions [88,89], an upregulation in older subjects may reflect a greater reliance on additional attentional resources [86]. As proprioception and sensory processing decline with age [90], the increased prefrontal involvement observed here may indicate compensatory engagement of attentional resources to support proprioceptive feedback processing.

Importantly, these changes of oscillatory properties could not only indicate functional recruitment but might also reflect adaptations in inhibitory neurophysiological mechanisms. Again, the timing and amplitude of alpha and beta oscillations are shaped by GABAergic inhibition, and stronger movement-related desynchronization has been linked to increased GABAergic activity [91,92,93] - a relationship that has also been demonstrated for TMS-evoked alpha- and beta-band desynchronization [94]. Thus, the observed prefrontal recruitment might as well arise from age-related alterations in inhibitory regulation within prefrontal circuits, reflecting a compensatory adaptation to declining sensorimotor efficiency. Overall, our results suggest a shift from an automatic, locally driven processing towards a more attention-dependent, top-down integration of sensorimotor information with advancing age.

## Conclusion

In conclusion, our study extends previous research on age-related adaptations in intracortical motor circuits, linking the HAROLD, PASA, and CRUNCH concepts to alterations in GABAergic inhibitory activity during aging. Importantly, while most studies on age-related changes in motor control have focused on task-based paradigms, our findings suggest that such mechanisms already apply in the absence of a cognitive or motor task. Specifically, our results suggest reduced GABAergic inhibition in the stimulated motor cortex, leading to diminished coupling with the surrounding and contralateral motor regions. In contrast, we observed upregulated activity in the prefrontal cortex, extending the findings of enhanced top-down control during aging to proprioceptive feedback processing. Together, these findings underline the relevance of alterations in inhibitory processes in the motor network in aging and suggest a shift from local, automatic towards a broader, more cognitively controlled sensorimotor processing in older individuals.

## Funding sources

This work was supported by the Research Training Group (RTG) 2783, funded by the German Research Foundation (DFG) - Project ID 456732630.

G.R.F., C.G., and C.T. are funded by the DFG – Project ID 431549029 - SFB 1451.

## Acknowledgements

We thank Luca Hladek for helpful discussions and insightful suggestions on figure design and title development. Moreover, we are grateful to Natascha Kellner for her technical assistance.

## Notes

### Competing Interest Statement

The authors have declared no competing interest.

### Author Declarations

Ethics committee of the medical faculty at the University of Cologne (file no. 17-244) gave ethical approval for this work.

