## Supplementary Material for "From Local Inhibition to Distributed Motor Control in Aging"

#### I. Motor hotspot and resting motor threshold (RMT)

We first identified the 'motor hotspot' of the primary motor cortex (M1), defined as the coil position eliciting motor evoked potentials (MEP) with the highest amplitude. Biphasic pulses were applied tangentially to the skull in a 45° posterior-anterior current direction, aiming to target the posterior wall of the precentral gyrus at the hand knob formation [1]. The EMG signal was measured using Ag/AgCl surface electrodes (Tyco Healthcare) from the first interosseous (FDI) muscle, then amplified, filtered (0.5Hz high pass and 30-300 Hz bandpass), and digitized using a Powerlab 26T device and the LabChart software package Version 8.0 (AD Instruments, Australia). Using an algorithm provided by the TMS Motor Threshold Assessment Tool (MTAT) 2.0 (http://www.clinicalresearcher.org/software.html)[2], we determined the resting motor threshold (RMT).

#### II. TMS-EEG recordings

A sampling frequency of 5kHz with a resolution of 0.1µV per bit was used to record the EEG signals. The data were filtered with a high-pass 0.1Hz and a low-pass 1kHz. Throughout the experiment, the impedance of all electrodes was kept below 5k$\Omega$ to ensure optimal signal quality. To record eye movements and blinks, we placed the two residual electrodes underneath the right eye and close to the outer corner of the left eye. Participants wore earplugs to reduce artifacts caused by the TMS click, such as auditory evoked potentials or eye muscle contractions [3,4]. Furthermore, we used a layer of plastic film between the TMS coil and the electrode cap to prevent bone conduction induced by TMS [5,6]. Since an increased coil-to-cortex distance might require a higher stimulator intensity to elicit an MEP, RMTs were reassessed after electrode cap placement. During the TMS-EEG recordings, subjects were asked to be at rest with eyes open. To account for the effect of hemispherical dominance, 14 of the 32 subjects (7 of the older subjects and 7 younger subjects) were stimulated in the left hemisphere. For all analyses, data of right hemispheric stimulation were flipped along the midsagittal plane, so that the stimulation site corresponded to the left hemisphere in all participants [7,8].

#### III. Data analysis

First, we visually examined the EEG signal to exclude single channels and trials contaminated by artifacts. Only data sets with at least 80 artifact-free trials per subject and less than 10 bad channels were used for further analyses to warrant sufficient quality (number of artifact-free single trials: younger subjects: 89.06 ± 4.9 SD, older subjects: 96.14 ± 13.43 SD; $p=0.01$, $Z=-2.51$, Mann-Whitney U-test). Data contaminated by the TMS artifact were removed in the time window 2ms before and 8ms after the TMS pulse and replaced with baseline extrapolations [9]. Subsequently, data were detrended, and a bandpass filter of 1-60Hz as well as a band-stop filter of 49-51Hz were applied (Butterworth 3rd order). The data were downsampled to 625Hz and segmented into windows ranging from –1000ms to +1000ms relative to the TMS pulse [10]. Subsequently, bad channels were spherically interpolated [11], and EEG signals were then average re-referenced and baseline-corrected [10]. Independent component analysis (ICA) was performed using the EEGLAB function *runica* to eliminate remaining TMS-related as well as ocular and muscle artifacts [12].

*Cluster-based permutation analysis*

Clusters were defined as two or more neighboring electrodes that demonstrated a t-statistic with an associated $p<0.05$. Identified clusters were then used for cluster-based analysis applying a permutation distribution generated with a Monte Carlo method (500 permutations) [13]. A cluster was compared to the permutation distribution and considered significant if the cluster-statistic (i.e., the sum of all t-statistics in each cluster) was smaller than the critical alpha value (two-sided, $p<0.05$).

### IV. Age-related alterations in TMS-evoked potentials after motor cortex stimulation

The cluster ranging from 90ms to 200ms post-stimulus involved electrodes in the ipsilateral centroparietal area surrounding the stimulation site ($p=0.008$, channels: C1, C2, C3, C5, CP1, CP2, CP3, CP5, CPz, Cz, P1, P2, P3, P5, PO3, PO7, Pz, T7). The second significant cluster from 60ms to 180ms comprised electrodes in frontal areas contralateral to the stimulation site ($p=0.012$, channels: AF3, AF4, AF7, AF8, F1, F2, F4, F6, F8, Fp2, Fpz, Fz, FC2, FC4, FC6).
